# Mortality reduction in 46 severe Covid-19 patients treated with hyperimmune plasma. A proof of concept single arm multicenter interventional trial

**DOI:** 10.1101/2020.05.26.20113373

**Authors:** Cesare Perotti, Fausto Baldanti, Raffaele Bruno, Claudia Del Fante, Elena Seminari, Salvatore Casari, Elena Percivalle, Claudia Glingani, Valeria Musella, Mirko Belliato, Martina Garuti, Federica Meloni, Marilena Frigato, Antonio Di Sabatino, Catherine Klersy, Giuseppe De Donno, Massimo Franchini

## Abstract

**BACKGROUND:** Hyperimmune plasma from Covid-19 convalescent is a potential treatment for severe Covid-19.

**METHODS:** We conducted a multicenter one arm proof of concept interventional study. Patients with Covid-19 disease with moderate-to-severe Acute Respiratory Distress Syndrome, elevated C-reactive Protein and need for mechanical ventilation and/or CPAP were enrolled. One to three 250-300 ml unit of hyperimmune plasma (neutralizing antibodies titer ≥1:160) were administered. Primary outcome was 7-days hospital mortality. Secondary outcomes were PaO2/FiO2, laboratory and radiologic changes, as well as weaning from mechanical ventilation and safety.

**RESULTS:** The study observed 46 patients from March, 25 to April, 21 2020. Patients were aged 63, 61% male, 30 on CPAP and 7 intubated. PaO2/FiO2 was 128 (SD 47). Symptoms and ARDS duration were 14 (SD 7) and 6 days (SD 3). Three patients (6.5%) died within 7 days. The upper one-sided 90%CI was 13.9%, allowing to reject the null hypothesis of a 15% mortality. PaO2/FiO2 increased by 112 units (95%CI 82 to142) in survivors, the chest radiogram severity decreased in 23% (95%CI 5% to 42%); CRP, Ferritin and LDH decreased by 60, 36 and 20% respectively. Weaning from CPAP was obtained in 26/30 patients and 3/7 were extubated. Five serious adverse events occurred in 4 patients (2 likely, 2 possible treatment related).

**CONCLUSIONS:** Hyperimmune plasma in Covid-19 shows promising benefits, to be confirmed in a randomized controlled trial. This *proof of concept* study could open to future developments including hyperimmune plasma banking, development of standardized pharmaceutical products and monoclonal antibodies.

## Introduction

Since the end of 2019, a new coronavirus strain has been reported in the Chinese province of Wuhan, indicated as 2019-nCoV or SARS-CoV-2^1-3^. The rapid spread of the epidemic in western countries has almost overcome the possibility of response of health systems leading to a high number of hospitalized people and deaths. There has been an inevitable lag between the onset of the pandemic and the availability of an effective therapy, and of today no treatment has been shown convincingly effective^4-7^. Previous data on the use of convalescent plasma (CP) during the SARS and MERS epidemics suggest the possibility of prompt immunization by administering specific antibodies using plasma from recovered/convalescent subjects ^8-16^. Also a metanalysis on hyperimmune immunoglobulins in severe acute respiratory infections of viral etiology, published in 2014, concluded for its efficacy and safety, though advocating well designed clinical trials^17^. To date there are very few studies in the literature that demonstrate the feasibility and efficacy of hyperimmune plasma in the SARS-CoV-2 pandemic ^18-20^.

For these reasons, we designed and conducted a *proof of concept* interventional multicenter study to show the potential efficacy and safety of hyperimmune plasma infusions, obtained from convalescent donors, in COVID-19 patients with respiratory failure, hospitalized at the participating Centers.

## Materials and methods

### Design

This is a *proof of concept* one-arm multicenter interventional study on the short-term (7 days) efficacy and safety of the infusion of hyperimmune plasma in moderate to severely compromised COVID-19 patients, according to Berlin Score. The evidence obtained will help either to plan an informed large clinical trial or to dismiss the proposed treatment if irrelevant. Primary endpoint of the study is 7-days mortality; secondary endpoints are respiratory function (PaO2/FiO2 ratio), laboratory (CRP, Ferritin, LDH, viral load) and radiologic changes, as well as weaning from mechanical ventilation (CPAP and or naso-tracheal intubation).

### Setting and population

Two university hospital and one general hospital in Northern Italy. This study has been registered at clinicaltrials.gov as NCT 04321421. It has been approved by the local Ethical Committee on March 17th, 2020 (Number n. 20200027967). Patients were enrolled from March 25, 2020 to April 21, 2020. Follow-up was closed on April 28, 2020.

Eligibility criteria are summarized in the eTable 1. Data were included into a database in REDCap hosted at the Fondazione IRCCS Policlinico San Matteo in Pavia and monitored remotely for missingness. The schedule of assessments is summarized in the eTable 2.

### Selection of convalescent donors and hyperimmune plasma

Male adults or female with no previous pregnancy COVID-19 recovered subjects, with 2 consecutive negative naso-pharyngeal swabs performed 7 to 30 days before, were recruited from the hospital records; their suitability was assessed according to the current Italian guidelines and transfusion law^21,22^. Plasma collection was performed with the latest generation cell separator (Trima Accel –Terumo BCT and Amicus – Fresenius Kabi) devices. A plasma volume of about 660 ml was collected during each procedure and immediately equally divided into two bags using a sterile tubing welder. Plasma pathogen reduction was performed with the INTERCEPT processing system (Cerus Europe BV) or the Mirasol PRT System (Terumo BCT, Lakewood, CO, USA). The collected units were stored at a controlled temperature ranging from −40 to - 25°C.^23^ (See supplementary material for further details).

### Plasma infusion

Plasma was delivered ready-for-use by the Immuno-Hematology Service to the Covid Units and was administered to the patients over 30 to 60 minutes, under supervision of the treating physician.

### Virology and immumunology

## SARS-CoV2 RNA detection

Total nucleic acids (DNA/RNA) were extracted from 200 ul of respiratory specimens. Clinical samples were pretreated with 1:1 ATL lysyis buffer and extracted using the QIAsymphony® instrument with QIAsymphony® DSP Virus/Pathogen Midi Kit (Complex 400 protocol) according to the manufacturer’s instructions (QIAGEN, Qiagen, Hilden, Germany). Specific real-time reverse transcriptase–polymerase chain reaction (RT-PCR) targeting RNA-dependent RNA polymerase and E genes were used to detect the presence of SARS-CoV-2 according to the WHO guidelines ^24^ and Corman et al. protocols^25^.

### SARS-CoV2 microneutralization assay

Neutralizing antibodies (NT-Abs) titers against SARS-CoV2 was defined according to the following protocol. Briefly, 50 μl of sample from each patient, starting from 1:10 in a serial fourfold dilution series, were added in two wells of a flat bottom tissue culture microtiter plate (COSTAR, Corning Incorporated, NY 14831, USA), mixed with an equal volume of 50 TCID_50_ of a SARS-CoV2 strain isolated from a symptomatic patient, previously titrated and incubated at 33°C in 5% CO_2_. All dilutions were made in EMEM with addition of 1% penicillin, streptomycin and glutammin and 5 y/mL of trypsin. After 1 h incubation at 33°C 5%CO_2_, VERO E6 cells [VERO C1008 (Vero 76, clone E6, Vero E6); ATCC^®^ CRL-1586™] were added to each well. At 48 h of incubation at 33°C 5%CO_2_wells were stained with Gram’s crystal violet solution (Merck KGaA, 64271 Damstadt, Germany) plus 5% formaldehyde 40% m/v (Carlo ErbaSpA, Arese (MI), Italy) for 30 min. Microtiter plates were then washed in running water. Wells were scored to evaluate the degree of cytopatic effect (CPE) compared to the virus control. Blue staining of wells indicated the presence of neutralizing antibodies. Neutralizing titer was the maximum dilution with the reduction of 90% of CPE. A positive titer was equal or greater than 1/10. Positive and negative controls were included in all test run.

### Sample size

Considering the hospitalization and mortality data retrieved from the Italian National Institute of Health by march 16^26,27^ we expect a mortality of about 15% (survival 85%; H0) in patients treated according to the standard of care. We also expect the mortality to decrease to 5% (survival 95%; H1) with the proposed hyperimmune plasma infusion. This being a proof of concept study, we use a one-sided type I error of 10%. According to the one stage Fleming design 43 patients will allow a power above 80% to reject H0. If we will observe at least 40 successes, the H0 hypothesis will be rejected and we might proceed with a future definitive trial. Three additional patients are enrolled to allow for possible drop-outs

### Statistical analysis

All continuous variables are summarized using the mean, standard deviation (SD) or the median, interquartile range (IQR). The frequency and percentage is reported for all categorical measures. All enrolled patients who receive the plasma infusion represent the analysis population.

#### Primary endpoint

The observed mortality rate is computed as the number of deaths over the full analysis population. The one-sided exact binomial confidence interval, at the 90% level (by design) is presented. The clinical and laboratory findings at baseline are described for patients surviving 7 days and listed for patients who died. No formal tests is performed.

#### Secondary endpoints

To assess changes in PaO2/FiO2, LDH, C reactive protein, Ferritin, and viral load over time we fit repeated measures linear (with Huber-White clustered robust standard errors to account for intra-patient correlation) or bootstrapped median regression models (depending on the distribution); the coefficient contrasting day 7 to day 1 together with its 95%CI are displayed to describe the end of study change.

## Results

### Study cohort

We enrolled 46 patients from 3 participating centers in this proof-of-concept study; patients were aged 63 years (SD 12) and 28 were male (61%). Mean oxygen saturation was 94% (SD 3) and the PaO2/FiO2 was 128 (SD 47); 14 (33%) had a severe Berlin score; 30 patients (70%) were on CPAP and 7 (16%) were intubated; 19 (41%) had 2 or more comorbidities and 38 (83%) had bilateral multilobe infiltrates at the chest radiogram. Patients had been having symptoms for 14 days (SD 7) and ARDS for 6 days (SD 3). More than 80% of patients were treated with antibiotics, hydroxychloroquine and anticoagulants (Table 1).

**Table 1:**
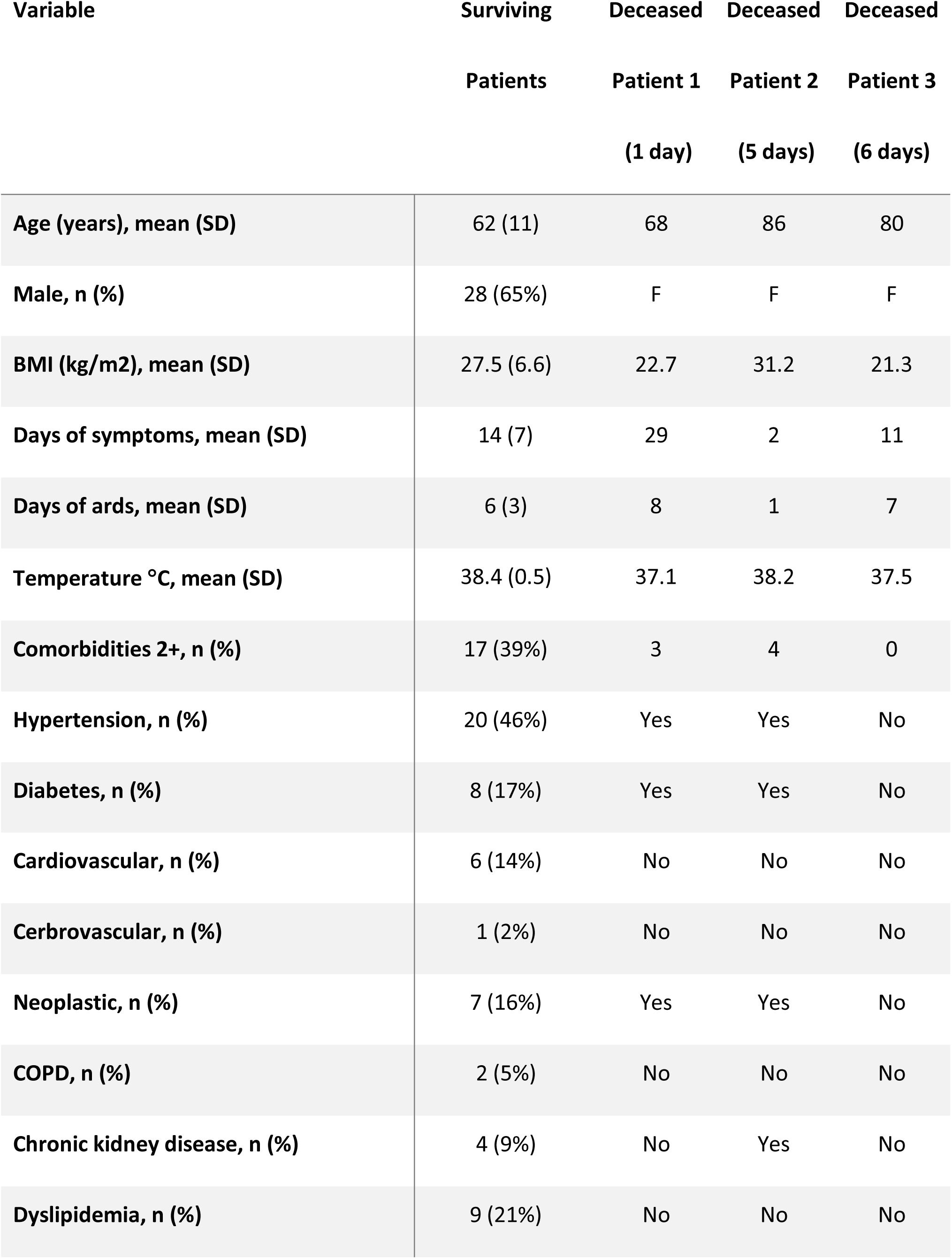
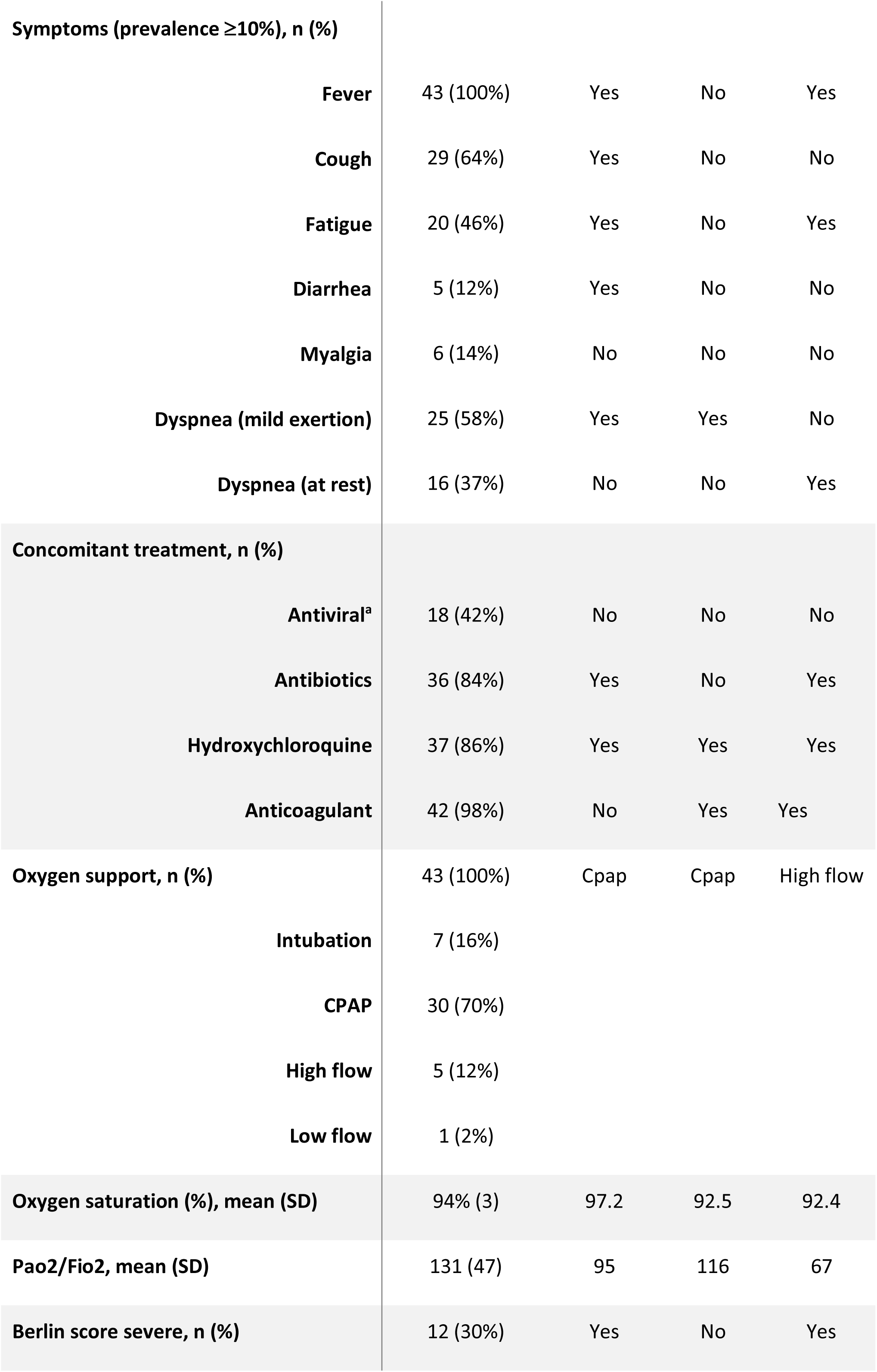
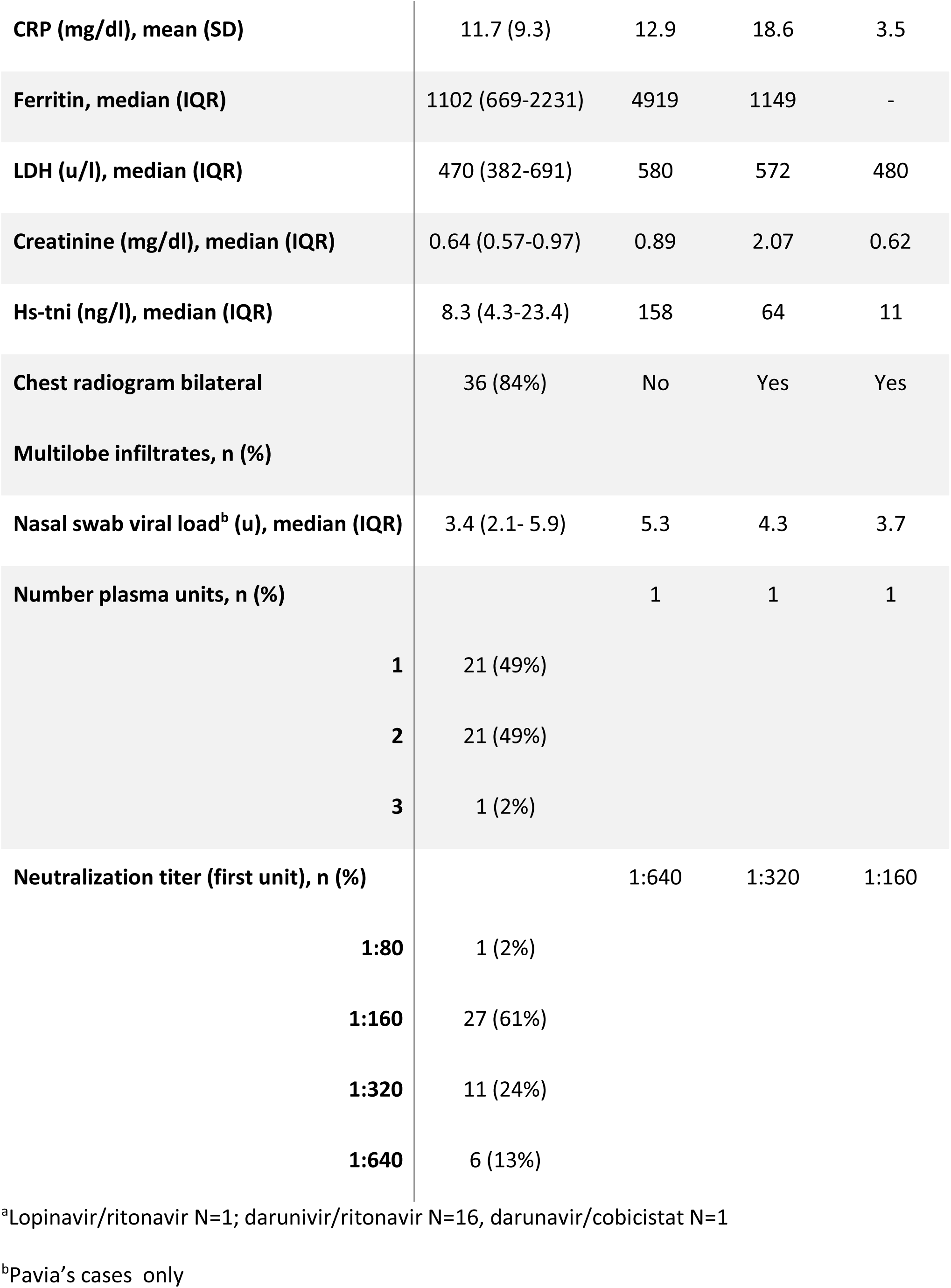
Description of medical history, baseline symptoms and laboratory findings

### Plasma Infusion

Twenty-four patients received one unit of plasma, 21 received two units and ne patient received 3 units. The plasma units administered at first infusion had a title of 1:160 or 1:320 in 85% of patients and of 1:320 in 12%; one patient only received 1:80 plasma (Table 1). At the second infusion titers were 1:80 in two patients, 1:160 in 11, 1:320 in seven and 1:640 in one. The third infusion performed in a single patient had a titer of 1:320.

The plasma infusion was well tolerated in 45/46 patients. It was interrupted in one case.

### Primary endpoint

Three patients out of 46 (6.5%) died within 7 days (at 1, 4 and 6 days); the upper one-sided 90%CI was 13.9% and 40 out of the first 43 patients enrolled had survived, allowing to reject the null hypothesis of a 15% mortality. The main characteristics of these 3 patients are listed in Table 1. Among them, two had important comorbidities, such as diabetes, hypertension and cancer, while the third had an extremely low PaO2/FiO2 level of 67 at the time of plasma infusion. Among survivors, the severity of the condition at baseline was confirmed by the low oxygen saturation (mean 94%) and PaO2/FiO2 (mean 131). More than 89% of patients showed bilateral multilobe infiltrates at chest radiogram and all laboratory biomarkers were markedly elevated (Table 1).

In a concurrent cohort of 23 patients from the Pavia Covid Registry, observed between March 10 and March 24 and satisfying the same entry criteria, the observed mortality was 30% (80%CI 18% - 46%).

### Secondary endpoints

At 7 days after plasma infusion PaO2/FiO2 increased by 112 units (95%CI 82 to142) in survivors, the chest radiogram severity decreased in 23% of patients (95%CI 5% to 42%); CRP, Ferritin and LDH all decreased by 60, 36 and 20%, respectively (Table 2 and Figure 1). Conversely no or little improvement was present in the three deceased patients (Figure 1).

**Table 2:** Changes from baseline in functional, laboratory and radiological parameters in survivors

| Variable | Day 1<br>N=43 | Day 7<br>N=43 | Difference and 95%CI |
| --- | --- | --- | --- |
| Oxygen saturation (%), mean (SD) | 94 (3) | 97 (2) | 3 (1 to 4) |
| Pao2/Fio2, mean (SD) | 131 (47) | 243 (40) | 112 (82 to 142) |
| Chest radiogram bilateral<br>Multilobe infiltrates, n (%) | 36 (84%) | 26 (61%) | -23% (-42% to -5%) |
| CRP (mg/dl), mean (SD) | 11.7 (9.3) | 4.7 (7) | -7.0 (-9.8 to -4.1) |
| Ferritin, median (IQR) | 1102 (669-2231) | 668 (406-1315) | -397 (-955 to 161) |
| LDH (u/l), median (IQR) | 470 (382-691) | 378 (323-422) | -92 (-157 to -27) |
| Viral load (U) ° | 3.4 (2.1- 5.9) | 0 (0-3.2) | -3.5 (-6.2 to -0.8) |

°Pavia's cases only

**Figure 1:**
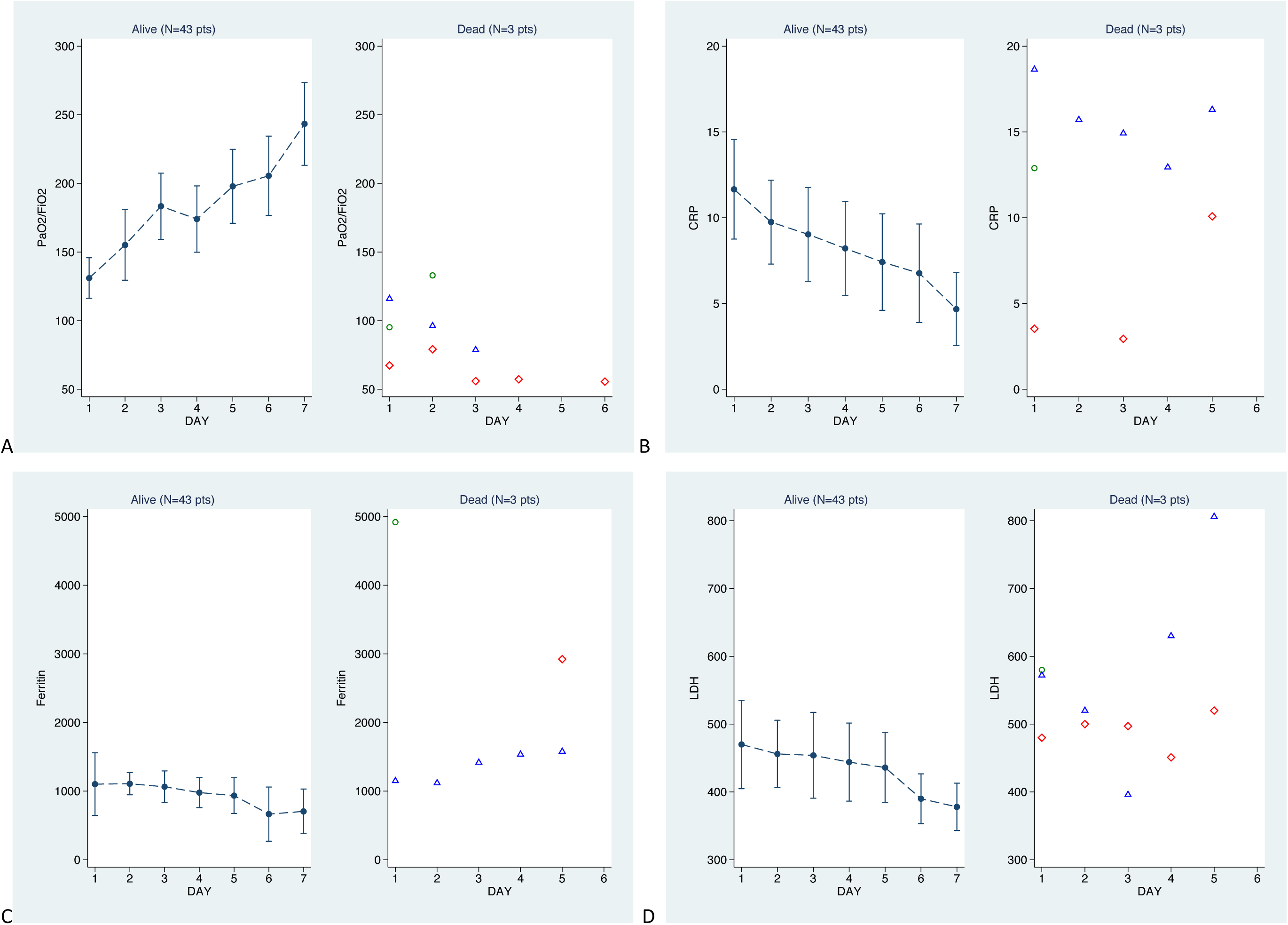
Changes over time of respiratory function and laboratory parameters from day 1 to day 7 for survivors and deceased patients. Panel A: PaO2/FiO2; Panel B: C reactive protein; Panel C: ferritin; Panel D: LDH Panel A & B: whiskers plots are mean and 95%CI; Panel C & D: whiskers plots are median and 95%CI

Overall 30 patients were on CPAP and 7 were intubated. Weaning from CPAP was obtained in 26 patients over a median time of 2 days (IQR 0-3) and 3 patients were extubated after a median of 2 days (IQR 1-5). Two out of 16 patients were discharged from ICU within 7 days from infusion, both at day 3.

Two patients were put on ECMO 1 and 6 days, respectively, after the plasma infusion.

### Safety

Five serious adverse events occurred in 4 patients. In one case the transfusion had to be interrupted. In two cases the relation to treatment was considered as likely and in two as possible (Table 3). Three adverse events had no spontaneous resolution and were treated accordingly.

**Table 3:**
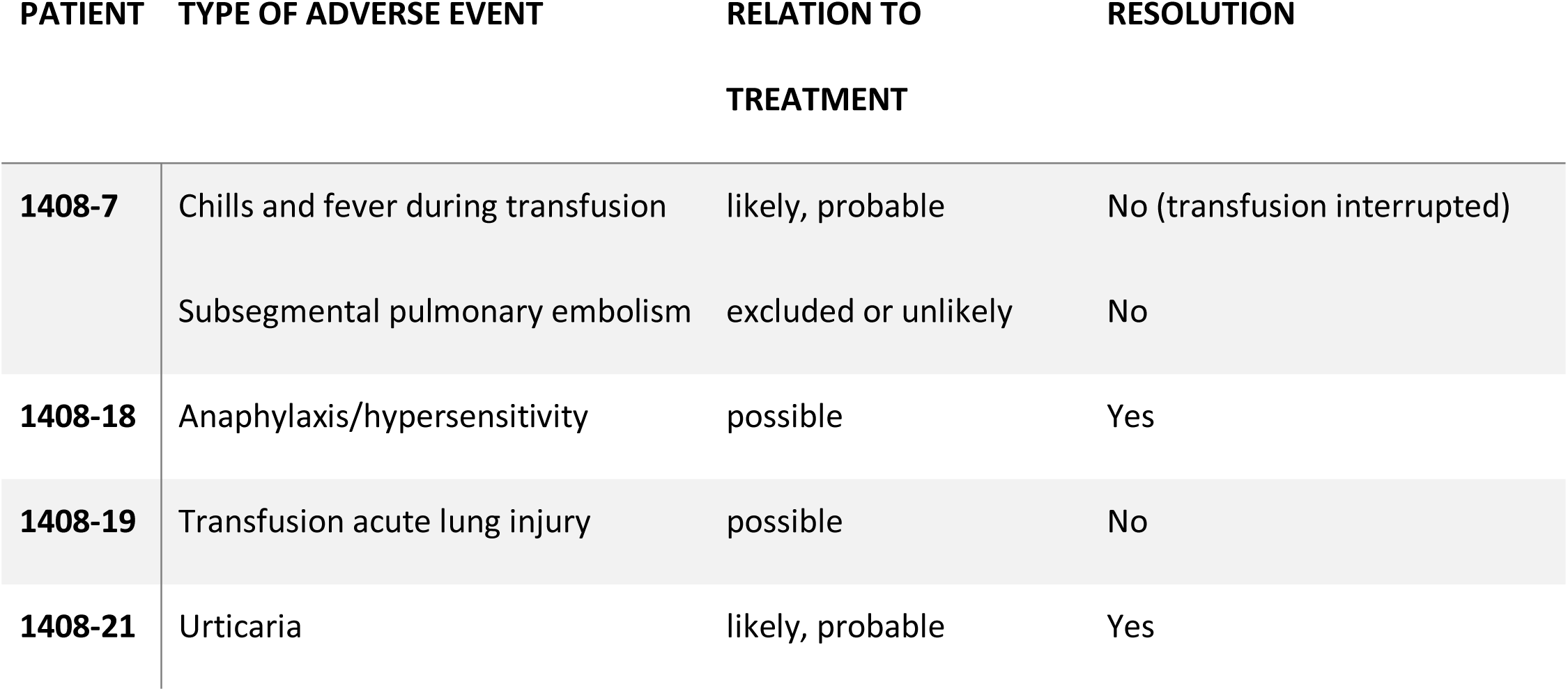
List of adverse events & relation to treatment per patient

| PATIENT | TYPE OF ADVERSE EVENT | RELATION TO<br>TREATMENT | RESOLUTION |
| --- | --- | --- | --- |
| 1408-7 | Chills and fever during transfusion | likely, probable | No (transfusion interrupted) |
|  | Subsegmental pulmonary embolism | excluded or unlikely | No |
| 1408-18 | Anaphylaxis/hypersensitivity | possible | Yes |
| 1408-19 | Transfusion acute lung injury | possible | No |
| 1408-21 | Urticaria | likely, probable | Yes |

## Discussion

This *proof of concept* study showed that the infusion of hyperimmune plasma in COVID-19 patients with a severe respiratory failure reduces short-term mortality by 2.5 times, from an expected 15% (that is about 1 out of 6 patients) at the time of study design to 6% (that is about 1 out of 15 patients). Compared with a concurrent series of 23 patients, satisfying the same entry criteria and observed in the period March 10^th^-24^th^, the decrease in the mortality rate was even more dramatic (5-fold, from 30% to 6%). Only three patients died during the seven days study period. Among them, two had important comorbidities, while the third had extremely low PaO2/FiO2 level at the time of plasma infusion.

Patients had been having symptoms for two weeks at the time of plasma infusion. Most had been on treatment with antibiotics, hydroxychloroquine and anticoagulants. In survivors, PaO2/FiO2 increased by two-fold, paralleled by a decrease in biomarkers: CRP by 60%, ferritin by 36%, and LDH by 20%. The bilateral multilobe infiltrates at chest radiogram disappeared in one-third of the study population. The viral load was reduced to null. Serious adverse events occurred in 4 patients; in two cases, they were likely related to the transfusion. Although hyperimmune plasma had been used for the treatment of severe cases in the 2002-2004 SARS outbreak ^8-14,17^, few data are available in the COVID-19 epidemics. Our results are consistent with these preliminary experiences from China. In a paper recently published in JAMA^18,19^ five critically ill patients at the Shenzhen Third People’s Hospital in Shenzhen, China, were treated with convalescent plasma at a neutralizing titer of 1:80 to 1:480, 10 to 20 days after admission. Clinical condition and laboratory findings had improved. As a result, three patients were discharged, and the other two were still hospitalized after one month. In a second study performed in three hospitals in Wuhan, China^20^ the outcomes obtained from ten patients with severe disease treated with convalescent plasma with high neutralizing titers (≥1:640), 16 days after the onset of symptoms were described. Following clinical and laboratory improvement, three patients had been discharged, and the other seven were ready for discharge. In contrast, in their historical controls, similar for baseline characteristics, only one patient improved, six were stable and three died. A third report of compassionate use of hyperimmune plasma gave encouraging results as well^28^.

On the contrary, in a fourth retrospective study^29^, administration of convalescent plasma in 6 patients led to suboptimal results. No previous determination of neutralizing antibody response had been performed. Indeed, both the FDA and the European Commission^30,31^ strongly recommend defined SARS-CoV-2 neutralizing antibody titers be measured in the donated plasma. While the FDA recommends a minimum neutralizing antibody titer of 1:160, though 1:80 might be acceptable, the EU considers titers of 1:320 or more to be optimal, though lower thresholds could be considered. In our study, of the 68 units of hyperimmune plasma administered, only three (4%) had a titer below 1:160; 58 (84%) had a titer between 1:160 and 1:320 and 7 had a titer of 1:640, largely consistent with the international recommendations. Of note, the promising efficacy of using neutralizing antibodies was described in a recent study that reported on a human monoclonal antibody neutralizing SARS-CoV-2 (and SARS-CoV) in cell culture^32^.

This study has some limitations, first of all the lack of a randomized control arm; it was however designed as a *proof of concept study* to verify the potential efficacy and safety of the administration of hyperimmune plasma in severely compromised Covid-19 patients and inform the design of a rigorous controlled randomized trial. However, the mortality was shown to be decreased by the treatment, both when compared to the mortality in Italy at the time of the design of hospitalized patients and to the mortality in our concurrent and similar observational cohort. Second, it was designed at the very beginning of the pandemic in Italy, and the patients were included under the pressure of medical emergency to provide them with an effective treatment at very short term. For this reason, some information was not planned to be collected, such as, but not only, the levels of D-dimer or other markers of inflammation, and long-term outcome.

In conclusion, we were able to show a promising benefit of hyperimmune plasma in Covid-19 patients, both through a reduction of mortality, an increase in the respiratory function and a decrease in the inflammatory indices. This is a *proof of concept* study, thus these findings should not be overinterpreted. Nevertheless, they pave the way to future developments including the rigorous demonstration of hyperimmune plasma efficacy in a randomized clinical trial, the need for hyperimmune plasma banking to anticipate a potential second wave of the pandemic, the development of standardized pharmaceutical products made from the purified antibody fraction (concentrated COVID-19 H-Ig) and, least but not last, the production of monoclonal antibodies on a large scale

## Data Availability

aggregated dta from corresponding author

## Authors contribution

Concept and design: perotti, baldanti, bruno, klersy, del fante

Acquisition, analysis or interpretation of data: all authors

Drafting the manuscript: klersy, del fante, baldanti, bruno, perotti, franchini

Critical revision of the manuscript for import ant intellectual content: all authors

Final approval of the version to be published: all authors

All authors agree to be accountable for all aspects of the work in ensuring that questions related to the accuracy or integrity of any part of the article are appropriately investigated and resolved.

## Access to the data

aggregated data (statistical analysis) are available from the corresponding author for metanalyses.

## Funding

none to declare

We thank Valeria Scotti, librarian at Fondazione IRCCS Policlinico San Matteo for her support on references. We also thank all the donors who after experiencing a difficult time being victims of the Covid-19 disease, are freely giving their convalescent plasma for the benefit of all.

## Covid-19 Plasma Task Force

| name | title | affiliation | contribution |
| --- | --- | --- | --- |
| Angelo Corsico | MD | Fondazione IRCCS Policlinico San Matteo, Pavia, Italy | patients enrollment and follow-up coordination |
| Federica Melazzini | MD | Fondazione IRCCS Policlinico San Matteo, Pavia, & University of Pavia Italy | patients enrollment and follow-up |
| Marco Lenti | MD | Fondazione IRCCS Policlinico San Matteo, Pavia, & University of Pavia Italy | patients enrollment and follow-up |
| Cristina Mortellaro | MD | Fondazione IRCCS Policlinico San Matteo, Pavia, Italy | donor's recruitment and data collection |
| Edoardo Vecchio<br>Nepita | MSc | Fondazione IRCCS Policlinico San Matteo, Pavia, Italy | virology laboratory analyses and data collection |
| Gianluca Viarengo | MD | Fondazione IRCCS Policlinico San Matteo, Pavia, Italy | donor's recruitment and data collection |
| Giorgio Iotti | MD | Fondazione IRCCS Policlinico San Matteo, Pavia, Italy | patients enrollment and follow-up coordination |
| Luciano Perotti | MD | Fondazione IRCCS Policlinico San Matteo, Pavia, Italy | patients enrollment and follow-up |
| Marco Maurelli | MD | Fondazione IRCCS Policlinico San Matteo, Pavia, Italy | patients enrollment and follow-up coordination |
| Margherita Sambo | MD | Fondazione IRCCS Policlinico San Matteo, Pavia, Italy | patients enrollment and follow-up |
| Mariangela Delliponti | MSc | Fondazione IRCCS Policlinico San Matteo, Pavia, Italy | Data collection and data entry |
| Raffaella Di Martino | MSc | Fondazione IRCCS Policlinico San Matteo, Pavia, Italy | virology laboratory analyses and data collection |
| Roberta Maserati | MSc | Fondazione IRCCS Policlinico San Matteo, Pavia, Italy | virology laboratory analyses and data collection |
| Valentina Zuccaro | MD | Fondazione IRCCS Policlinico San Matteo, Pavia, Italy | patients enrollment and follow-up |
| Gennaro Mascaro | MD | Ospedale Maggiore della Carità, Novara, Italy | patients enrollment and follow-up |

## SUPPLEMENTARY MATERIAL

**Table of contents**

eTable 1

eTable 2

Plasma collection from the selected donors and validation procedures

Consort checklist

**eTable 1.** - Eligibility criteria.

| Inclusion criteria |  |
| --- | --- |
| 1 | Age $\geq 18$ years |
| 2 | Positive SARS-CoV-2 RT-PCR on nasal swab or deep respiratory sample |
| 3 | Diagnosed with moderate-to-severe ARDS (Acute Respiratory Distress Syndrome) for $\leq 10$ days, according to Berlin definition:<br>a. New or worsening of respiratory symptoms within a week of a known clinical insult;<br>b. Radiological imaging (CT, RX, Ultrasound) of bilateral pulmonary opacities not fully explained by effusion, lobar or pulmonary atelectasis, or nodules;<br>c. Respiratory failure not fully explained by heart failure or fluid retention<br>d. $\text{PaO}_2 / \text{FiO}_2 \leq 200$ mmHg with PEEP (or CPAP) $\geq 5$ cmH<sub>2</sub>O |
| 4 | Increase in the PCR value of approximately 3.5 times the upper reference limit or above 1.8 mg / dl |
| 5 | Need for mechanical ventilation and / or CPAP |
| 6 | Patients who signed the informed consent. If there is no possibility of obtaining informed consent for the clinical condition (e.g. patients sedated and treated for acute respiratory failure and consequent mechanical ventilation), the patient's consent will be assumed until manifestly stated otherwise. |
| Exclusion criteria |  |
| 1 | Diagnosis of moderate-severe ARDS for $> 10$ days |
| 2 | Patients with proven hypersensitivity or allergic reaction to blood products or immunoglobulins |
| 3 | Manifest unwillingness to participate |

**eTable 2.** - Schedule of assessments.

| **Study Period:** | **Screening** | **Day 1**<br>**(baseline)** | **Day**<br>**2** | **Day**<br>**3** | **Day**<br>**4** | **Day**<br>**5** | **Day**<br>**6** | **Day**<br>**7** | **End of**<br>**Study** |
| --- | --- | --- | --- | --- | --- | --- | --- | --- | --- |
| Informed Consent | (X) |  |  |  |  |  |  |  | (X) |
| Inclusion/Exclusion Criteria | X |  |  |  |  |  |  |  |  |
| Demographics | X |  |  |  |  |  |  |  |  |
| Medical History | X |  |  |  |  |  |  |  |  |
| Physical examination |  | X | X | X | X | X | X | X |  |
| Laboratory |  | X | X | X | X | X | X | X |  |
| Hemogas analysis |  | X | X | X | X | X | X |  |  |
| Viral load (nasal swab sputum, BAL) |  | X |  | X |  |  |  | X |  |
| Chest X rays |  | X |  | X |  |  |  | X |  |
| Oxygen Support |  | X | X | X | X | X | X | X |  |
| Concomitant treatments |  | X | X | X | X | X | X | X |  |
| **COVID-treatment (PLASMA)** |  | **X** |  | **(X)°** |  | **(X)°** |  |  |  |
| Outcomes |  |  |  |  |  |  |  |  | X |
| Adverse events |  | X | X | X | X | X | X | X |  |
| immune response |  | X | X |  |  |  |  |  |  |
| Blood sample collection for storage<br>(exploratory biomarkers in future) |  | X |  | X |  |  |  | X |  |

### Plasma collection from the selected donors and validation procedures

Donors were male or females with no previous pregnancies, aged 18 or above, who had recovered from Covid-19 disease (defined as 2 consecutive negative naso-pharingeal swabs) since not less than 7 days and not more than 30 days. The donors were registered according to the national regulation and thoroughly clinically evaluated by the local physician, with the purpose of highlighting any absolute contraindications to the aphaeresis procedure. All donors will need to test negative for hepatitis A and E RNA, and parvo virus 19 DNA, as well as for hepatitis B, C, HIV and syphilis at the molecular test (according to the current law). All convalescent patients were pre-tested (72 hours in advance) for anti-SARS-CoV-2 neutralizing antibodies title except those living far from the hospital which were tested at the time of donation.

Plasma collection was performed in a dedicated facility, using latest generation cell separator (Trima Accel - Terumo BCT and Amicus-Fresenius Kabi) devices, set according to the donor characteristics, under nurses’ supervision. A plasma volume of about 660 ml was collected during each procedure and immediately divided in two bags of equal volume, using a sterile tubing welder. Then, plasma pathogen reduction was performed with the INTERCEPT processing system (Cerus Europe BV) or the Mirasol PRT System (Terumo BCT, Lakewood, CO, USA), as specifically required by the National Centre for Blood and labelled as hyperimmune Covid plasma. Finally, it was stored in a dedicated freezer, at a controlled temperature ranging from −40 to −25°C. Collected plasma had a neutralizing title of 1:160 or more.

As per routine, the plasma was validated and made available for infusion at the completion of all tests.

Request of ABO compatible plasma was performed by treating physician using the established local procedures, inclusive of electronic tracking.

